# A Model of Workflow in the Hospital During a Pandemic to Assist Management

**DOI:** 10.1101/2020.04.28.20083154

**Authors:** Marc Garbey, Guillaume Joerger, Shannon Furr, Vid Fikfak

**Affiliations:** The Houston Methodist Research Institute, Weill Cornell Medical College, Houston, TX, USA; ORintelligence, Houston, TX, USA; LaSIE, UMR CNRS 7356, University of la Rochelle, France; Department of Surgery, University of Texas, Texas Health Sciences Center, El Paso, TX, USA

## Abstract

We present a computational model of workflow in the hospital during a pandemic. The objective is to assist management in anticipating the load of each care unit, such as the ICU, or ordering supplies, such as personal protective equipment, but also to retrieve key parameters that measure the performance of the health system facing a new crisis. The model was fitted with good accuracy to France’s data set that gives information on hospitalized patients and is provided online by the French government. The goal of this work is both practical in offering hospital management a tool to deal with the present crisis of COVID-19 and offering a conceptual illustration of the benefit of computational science during a pandemic.

## Introduction

During a pandemic, such as the one of the coronavirus disease 2019 COVID-19, management of patient flow and hospital resources are pushed to their limits: Hospital Emergency Treatment Facilities through intermediate care unit (IMU) and intensive care unit (ICU) are or are going to be be severely strained. Healthcare professionals are often times forced to make difficult decisions in patient care and resource allocation. Patient profiles might be out of the ordinary routine of the hospital and workflow must be different. End-to-end on demand visibility with identification of real constraints is needed for the senior management.

A manager may have simple but essential questions such as: how many beds do I need on the floor, how many beds are available in the critical care unit, how much supplies should be ordered to take care of our patients and protect our staff from infection, how long will the facility have to work at maximum capacity, is there enough staff to hold this workload long enough, are we doing well with patient outcomes, etc.

Multiple governmental and private agencies have focused on creating dashboards for easy access and understanding of global pandemic data. These applications are great to give the general assessment of the pandemic but do not allow a projection of detailed information at the local hospital scale that is necessary to optimize the management of patient workflow.

There is a significant amount of literature about mathematical models in epidemiology that provide a rigorous framework to make predictions on the number of people who are going to be symptomatic enough to require hospitalization. This approach has been quickly applied to COVID-19 with success [20, 25]. In the case of the COVID-19 pandemic, it is particularly difficult because large bodies of infected people are asymptomatic. Consequently, the basic reproduction number *R*_0_ factor of COVID-19 is still under active debate.

On the hospital workflow side, while there is a large amount of work on this topic [23], one of the difficulties is to asses the death rate of patients hospitalized at the beginning of the pandemic because the Length of stay (LOS) is rather long and the disease is still not well understood [2, 16, 26]. Every hospital has to adapt to the new crisis as it arrives, so clinical practice may vary greatly from one institution to another. A number of guidelines and great reports have been quickly edited to support the heath community, but it takes time to standardize the healthcare process [3, 5, 13, 24].

Our goal in the paper was to come up with a simple and robust mathematical framework that is easy to use and that supports the management of the patient workflow during a pandemic. Such a model should operate on a relatively limited data set that reports daily on the number of patients admitted for hospitalization, patient output of the facility (such as number of patients healed per day or number of deaths per day), and at minimum the number of patients staying in the most critical unit of the facility - the ICU. The model can then be customized to the local hospital system with an optimization technique to achieve calibration after a few weeks of data acquisition. Much more can be done with the patient electronic records that detail patient comorbidities and chronic conditions, provided that the disease of the pandemic is well understood.

We have used a Markov process description of the workflow’s graph with probability governing the patient transition from one care unit to another, as well as a simple statistical model of patient LOS at each stage. We will show that with a minimum number of parameters used to fit on the time series listed above for a period of a few weeks, one may start to assemble the information needed to assist the senior management in getting answers and identifying real constraints to reduce speculation or misallocation of resources.

This work is our first iteration to achieve a very ambitious goal: as data becomes available, the quality and level of detail of modeling should keep improving to achieve better results. It is our hope that such an effort, among many others, will once again prove how much digital health can benefit from computational science to improve patient care.

The paper is organized as follows: Section 2 describes our method to construct the model and details the choices we made to work with the data set on hand; Section 3 gives the main results and solution to our initial goal in supporting management; Section 4 discusses the benefit and limitation of our method and concludes with further potential development.

## Materials and methods

Because of the sparsity of data available to construct a predictive model during a pandemic crisis, we are going to use a very simple model that reproduces the workflow of Table 1. Let’s start with a brief description of standard patient workflow -see Figure 1 -with respect to disease progression -see Figure 2.

**Table 1.**
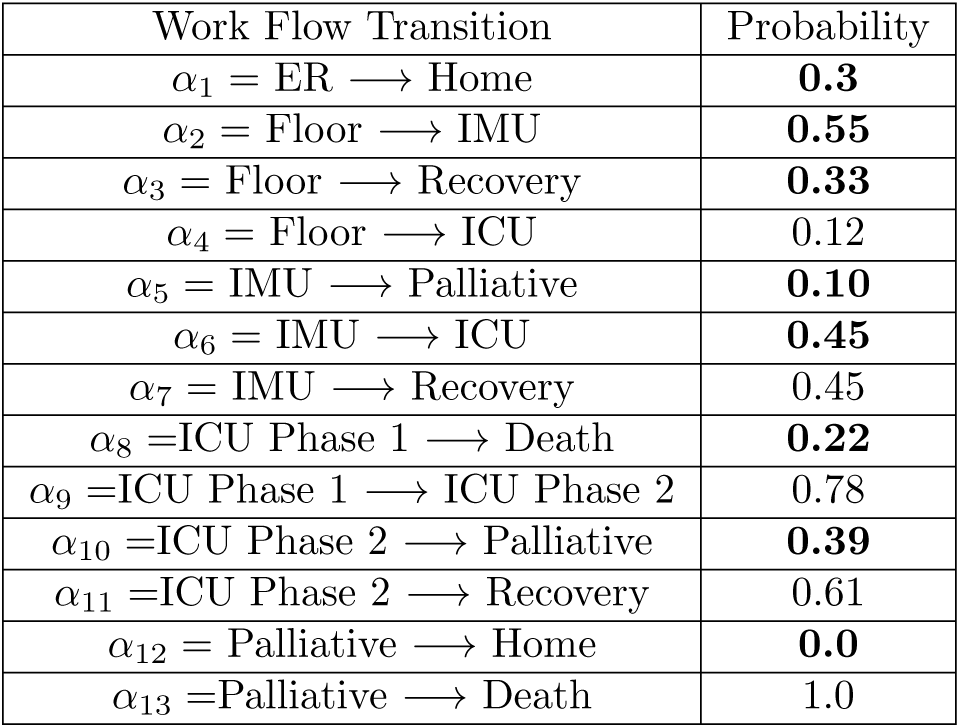
Probability of Transition for the Patient in reference to the Workflow of Figure 1.

| Work Flow Transition | Probability |
| --- | --- |
| $\alpha_1 = \text{ER} \rightarrow \text{Home}$ | **0.3** |
| $\alpha_2 = \text{Floor} \rightarrow \text{IMU}$ | **0.55** |
| $\alpha_3 = \text{Floor} \rightarrow \text{Recovery}$ | **0.33** |
| $\alpha_4 = \text{Floor} \rightarrow \text{ICU}$ | 0.12 |
| $\alpha_5 = \text{IMU} \rightarrow \text{Palliative}$ | **0.10** |
| $\alpha_6 = \text{IMU} \rightarrow \text{ICU}$ | **0.45** |
| $\alpha_7 = \text{IMU} \rightarrow \text{Recovery}$ | 0.45 |
| $\alpha_8 = \text{ICU Phase 1} \rightarrow \text{Death}$ | **0.22** |
| $\alpha_9 = \text{ICU Phase 1} \rightarrow \text{ICU Phase 2}$ | 0.78 |
| $\alpha_{10} = \text{ICU Phase 2} \rightarrow \text{Palliative}$ | **0.39** |
| $\alpha_{11} = \text{ICU Phase 2} \rightarrow \text{Recovery}$ | 0.61 |
| $\alpha_{12} = \text{Palliative} \rightarrow \text{Home}$ | **0.0** |
| $\alpha_{13} = \text{Palliative} \rightarrow \text{Death}$ | 1.0 |

**Fig 1.**
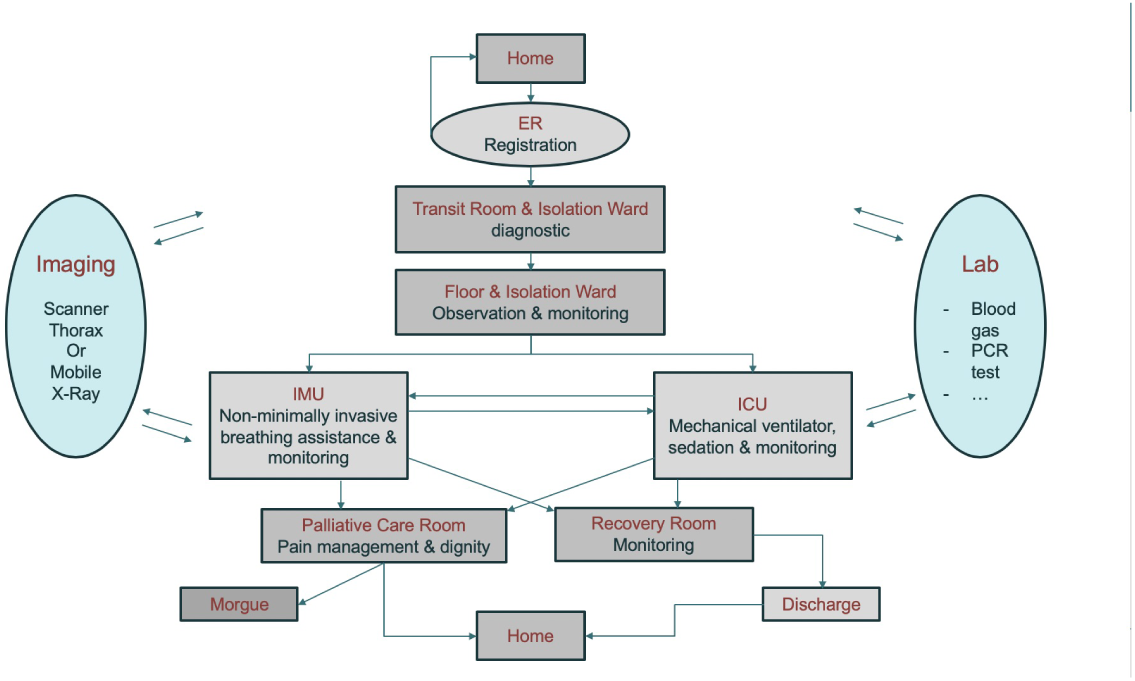
Workflow used in the model to follow patient progression in the hospital.

**Fig 2.**
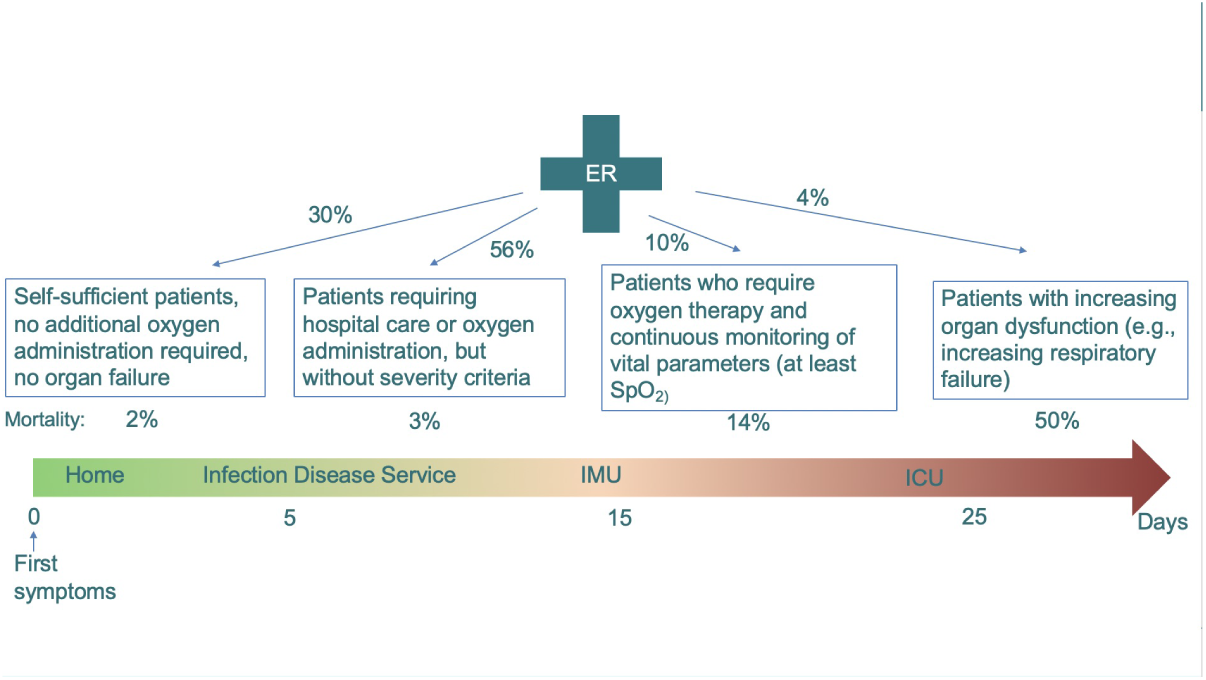
Disease Progression -see references [8, 17].

The patient moves from one care unit to another according to his/her condition. The first two steps are registration and diagnostics, which in principle should be a relatively quick process. For the patients who stay in the hospital because their health condition justifies a longer stay, they are first put in a ward unit for further assessment and treatment. This step is where a number of medical imaging steps start involving either a chest CT scan in the imaging center or a chest X-ray with a mobile unit. Meanwhile, significant biological lab work starts to grade the patient’s condition more precisely and continues during the patient’s stay. These resources, i.e. imaging and lab work, are typically shared by all patients in the hospital and therefore may slow down the process. For simplicity and in the absence of adequate data set for validation, we neglect these constraints. Some of the patients who receive medical attention do well with conservative management only and can be discharged home after a few days. But for others, their health condition may deteriorate and those patients will need to be moved to the IMU for higher level of care and/or to transfer to the ICU for ongoing monitoring and mechanical ventilation. The IMU and ICU require extensive supplies and resources. It is often mentioned that the number of available ventilators is critical to ICU functions. However, it is not the only limiting factor: patients under mechanical ventilation need sedation and might be connected to a number of additional systems to deal with organ failures. Once again for simplicity and because of the lack of input data, our model will not take into account these bottlenecks. There are no technical difficulties required to add those constraints in the mathematical model with our bottom up description of the workflow as in [10, 15]. Additional steps can be recovery for patient being well or unfortunately palliative care when the patient is not responsive to treatment. There are many exceptions and singularities to these standard paths: for example, a patient may go directly from admission to the ICU when their condition is too unstable. In some hospitals, the floor might be shared by patients who are recovering from COVID-19 and palliative care patients. Despite this, we will separate these functional units in our model to clarify the workflow process according to what each patient stage requires in terms of resources and time to deliver adequate care. To summarize, a simple workflow graph is created and the main requirement is to know (i) the probability that a patient goes from one care unit to another and (ii) a statistical estimate of how long the patient should stay in each care unit before moving on.

Our model follows a Markov process for (i): there is a probability associated with each branch of the graph summarized in Table 1. With respect to (ii), we use a lognormal distribution that can be reconstructed from the parameters listed in Table 2 and Table 3. This simple framework allows us to construct a generic model that dynamically computes the number of patients for each care unit as a function of the number of patients showing up in the ER. In particular, we get a time series of the number of patients staying in the ICU, which is the most critical care unit in term of resource allocation, as well as the number of patient outputs, such as the number of patient healed and discharged per day, or the number of death(s). These time series can be fitted to existing data the hospital obtains during a period of a few weeks prior to retrieving the performance parameters of Table 1. Once the model is calibrated, it can be used to extrapolate the load of each care unit in the next few days and anticipate the need of staff and supplies -see Table 4.

**Table 2.**
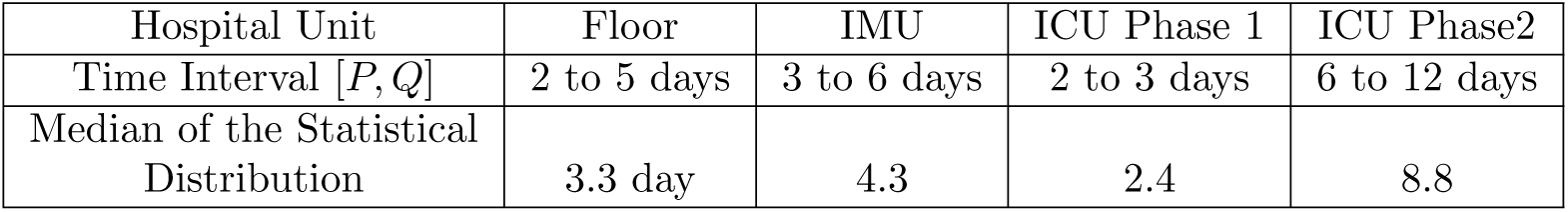
Time Window for the Patient Stay at each Stage in reference to the Workflow of Figure 1.

| Hospital Unit | Floor | IMU | ICU Phase 1 | ICU Phase2 |
| --- | --- | --- | --- | --- |
| Time Interval $[P, Q]$ | 2 to 5 days | 3 to 6 days | 2 to 3 days | 6 to 12 days |
| Median of the Statistical Distribution | 3.3 day | 4.3 | 2.4 | 8.8 |

**Table 3.** Time Window for the Patient Stay at each Stage in reference to the Workflow of Figure 1.

| Hospital Unit | Registration | Diagnostic | Recovery | Palliative | Discharge |
| --- | --- | --- | --- | --- | --- |
| Time Interval $[P, Q]$ | 15 to 50mn | 3 to 9h | 1.5 to 4 days | 0.5 to 2.5 days | 30mn to 3h |
| Median of Statistical Distribution | 35mn | 5.6h | 2.5 day | 1.5 day | 1.5h |

**Table 4.** Number of Staff required at each care unit per beds in reference to the Workflow of Figure 1.

|  | Floor | IMU | ICU | Recovery | Palliative |
| --- | --- | --- | --- | --- | --- |
| Nurse | 5 beds | 2 beds | 2 beds | 4 beds | 3 beds |
| MD | 10 beds | 10 beds | 6 beds | 10 beds | 10 beds |

This discrete model is stochastic, so one needs to run many simulations to build a statistical estimate of such quantities. It is appropriate to retrieve the unknown parameters of the model using a form of stochastic optimization method, such as genetic algorithm, since the model workflow process, like the one in the hospital, is discrete, noisy, and nonlinear.

Let us describe the data set we are using to construct our model. The French government has kindly decided to release the records of most public hospitals around the country during the COVID19 crisis. From this excel file, we can easily recover the number of patients staying in hospitals, the number of patients in ICU, the number of patients healed and discharged, and the number of patients dying in a medical institution. Those numbers are updated daily and go back to March 18, 2020 [27]. We will extensively use this French Data Set (FDS) to identify the missing parameters of our model.

The number of parameters of our model is relatively large: about one parameter for each branch of the graph minus the number of nodes for (i) and two parameters for the log distribution of (ii) in each care unit. To avoid over-fitting, one should come up with a strategy that lowers the number of unknown based either on literature or hypothesis that can be validated otherwise. We are going to describe thereafter the rationale for our choices to the best of our knowledge and further discuss some of the limitations of our model in Section 4.

First of all, a lognormal distribution of the duration of each step of the process might be justified as follows. Biological process, such as incubation and recovery, are often described as such [18, 19]. First, the patient’s condition is indeed dominated by his/her biological time. Second, medical procedures with their associated time lag and delay are also often best described as lognormal processes [10, 22] with a long tail. This is not in contradiction with the fact that patient LOS in the hospital may not ideally be described by a simple exponential distribution or similar. Overall, LOS adds up the time distribution of each step in a Markov process and might be described at the convolution of the probability distribution of each step [14].

Now let’s review the parameters of Table 1 that gives the probability transition from one unit to another, in order to rationalize the construction of our generic model. One can first list the following constraints assuming that all possible paths are exhaustively listed in the workflow of Figure 1, so we have:

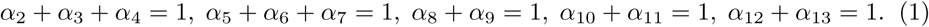

Overall, the death rate and recovery rate of patients who are staying in the hospital should be within an acceptable limit. Technically, the death rate of hospitalized patients is:

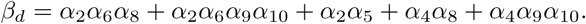

Similarly, the recovery rate of hospitalized patients *β_h_* =1 − *β_d_* is:

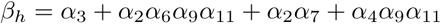

*β_d_* is difficult to asses with a pandemic that just started. As a matter of fact, most infected patients are still in the hospital and their outcome may not be clear. We look thereafter for some lower and upper bounds of *β_d_* that limits our search.

According to the Intensive Care National Audit & Research Center (ICNARC) report of March 27, 2020 [28], we may conclude that a lower bound to overall death of patients admitted into ICU units is about 10% - as a matter of fact most patients were still in the hospital at this early stage. On the other hand, and based on a very small case series in the Seattle region [1], the death rate was up to 50%, but in this study most patients had chronic medical conditions. These two publications illustrate the difficulty of recovering the true rate of death for today’s large population whom have been hospitalized, mostly due to the heterogeneity of the population they encompass.

In France, as of April 17, 2019, the number of deaths in hospitals was 11,842 patients and recovered was 35,983 patients [27]. Assuming that the proportion of death versus recovery will be about the same for the patients who are still ill, the death rate of hospitalized patients should be around 25%. Finally, according to [26], an early estimate of the death rate for hospitalized patients in Wuhan, China based on a case series of 191 patients was 54*/*191 = 28%.

We restrict ourselves to the model matching the FDS to a [10%, 40%] death rate interval, that is:

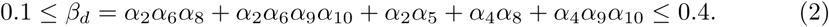

According to several reports including the ICNARC one mentioned above, it is expected that the number of patients dying in ICU is about 50%. [6] provides much further details on the probability of survival of patients with ARDS under mechanical ventilator as a function of the day of the start. It shows that about 25% of the patients in ICU die during the first few days from severe complications. We will introduce an artificial two phases ICU decomposition of the patient stay in the ICU to bypass the limitation of a single lognormal distribution that may not represent an adequate model of LOS in this unit according to [6] clinical studies: a short phase one with mortality driven by *α*_8_ and a longer phase 2 with mortality driven by *α*_10_.

Consequently, we will assume that:

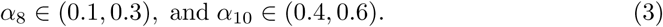

There are also few parameters in Table 1 that should have near to no limited effect on statistics when matching our model to FDS. FDS is based on hospitalized patients, so *α*_1_ cannot be recovered from this data set. According to FDS, about 30% of patients who show up at the emergency room (ER) are returning home [27]. We will choose *α*_1_ =0.3.

According to Dr. M. Mueller [29], 25% of the patients who are not responsive to treatment may leave palliative care alive and are discharged home. This may vary depending on each country or hospital policy. Because patients with COVID-19 in palliative care are still very contagious, we will assume they stay in the hospital until the end. We will choose *α*_12_ =0., for all our calculations.

To sum up, our model essentially needs the calibration of 6 parameters, namely

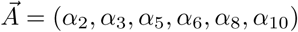

under the set of constraints (1), (2), (3).

Let us denote *F^admission^*(*jd*) the number of patients admitted per day *jd* ∈ 1*..N* in the hospital who have a positive diagnosis and must stay in the hospital. We will use this time series as an input to our model simulation in order to calibrate the model against the FDS.

Let us denote 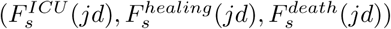 the time series of patients in the ICU, patients healed and discharged home, and patients who died per day *jd* ∈ 1..*N* obtained from the simulation output. We will denote respectively 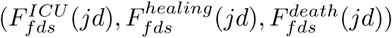 the times series extracted from the FDS.

We find 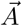 as the solution of the minimization problem of the weighted norm:

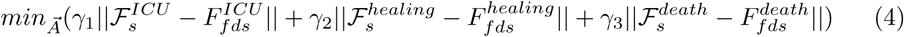

where 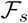 is the mean of a large number of runs of the model. This number of runs is set large enough to let the solution of the optimization problem be independent of it. As mentioned above, we will use a genetic algorithm to solve that minimization problem. The weight factor (*γ*_1_*,γ*_2_*,γ*_3_) in (4) can be set equal or unequal to favor the quality of the fitting for one of the variables, such as the number of patients in the ICU that is critical to management.

Table 2 and 3 give the time window we used for each transient stage. We construct a lognormal distribution of duration for the patient stay in such a way that about 90% of the patients’ stay will be within a coarse approximation [*P, Q*] listed in these tables. The choice of the parameters in Table 2 and Table 3 might be easier to come up with.

One of the most remarkable features is that patients with COVID-19 who stay in the ICU can be longer than usual [1]. The LOS in palliative care was set according to Dr. M. Mueller’s data [29]. We have used extensively [2,26], as well as the feedback from clinicians in the field to estimate the interval of variation for the parameters [*P*, *Q*] the best we could. We used a fairly large interval since it can be observed that the standard deviation for LOS in each care unit is large as described in this report from the Imperial College London COVID-19 Response Team [30]. One may fine tune the interval value [*P*, *Q*] if needed in the fitting process of the model to the data set of time series available.

To distinguish those unknown parameters that are important from those who are less significant, we run linear sensitivity analysis for each of our results. This method is used to confirm that the time window parameters of Table 2 and Table 3 have a secondary effect on the quality of the model fitting.

Finally, we derive from our model some predictions on staffing and supplies for the next week or so, as well as the load foreseen for each care unit. The nature of the stochastic simulation automatically gives an uncertainty estimate on these predictions that increases as time grows. To compute supplies such as personal protection kits, we can use some adaptation of the reference of the CDC web site [31] that was constructed for Ebola. Our software can then be used to feed the stock management scheme implemented by CDC for COVID-19 [32]. In this paper, we use a growth estimate of two personal protective equipment (PPE) per shift and per staff member for simplicity.

Similarly, we have listed in Table 4 a gross approximation of the number of nurses and staff per bed site in each unit. Those figures are depending on the crisis situation and might differ depending on the country [5].

In order to take into account the fact that staff and supplies are limited and require hard management choices during a pandemic crisis, we tested the model further against the scenario of a shortage on nurses who are essential in intensive care units. To introduce a risk factor due to the shortage of nurses, we have extrapolated from [9] and [8], a scenario where a 40% shortage of nurses results in:

- Time spent on floor increases by 15%
- Chances of transition from floor to Recovery or floor to IMU decreases by 15%
- Chance of dying in ICU increases by 15%

To get a continuous approximation, we assume that the shortage of nurses has a linear effect, and use linear interpolation for shortages from 0% to 40% maximum. This is certainly a gross approximation, but we felt that it was important to bring awareness to those effects with a simulation tool. We will present in the next section our results.

## Results

Let us first report on the model fitting with the FDS. We sum up the number of admissions, patients in ICU, number of recoveries and deaths for the whole country of France in order to get a robust data set that averages the noise of the data. We calibrated the model to this largest data set that covers the period 3/18/20 to 4/11/20 and found a death rate of about 25%. This result is in agreement with the estimate we did in Section 2, as of April 17, 2019 [27].

Figure 4 and 5 show the results with the parameters as listed in Table 1 to 3. All numbers have been scaled by a factor to represent an average hospital size. The sensitivity analysis on the alpha unknown vector 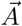 is reported in Figure 3.

**Fig 3.**
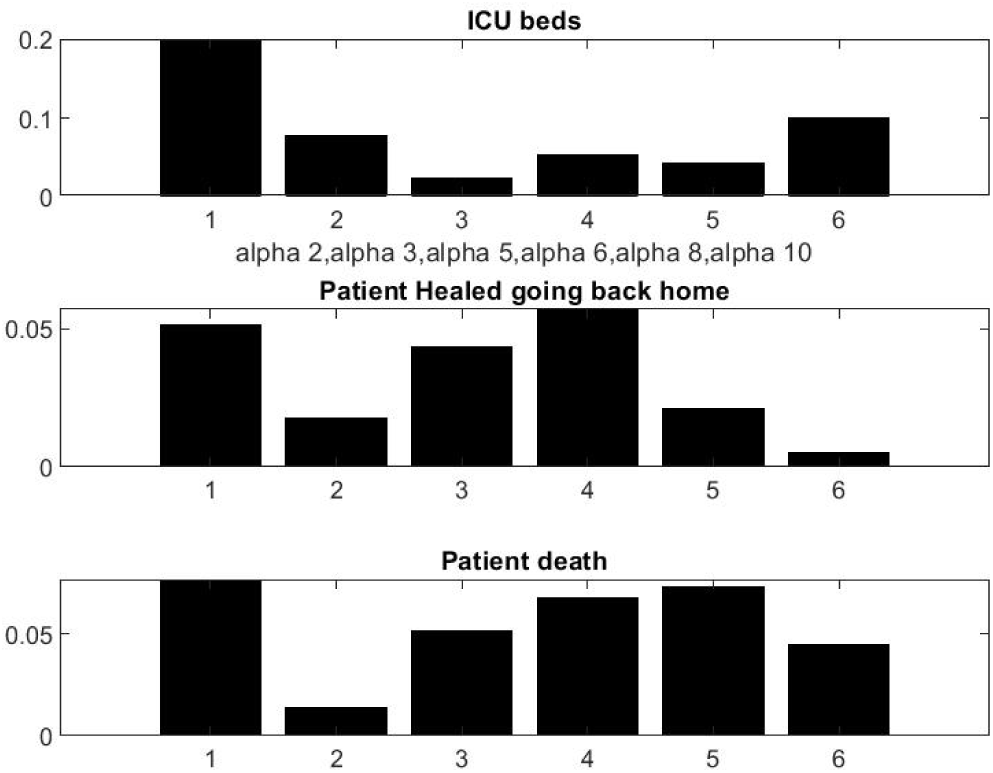
Linear sensitivity with respect to parameters [*α*_2_*,α*_3_*,α*_5_*,α*_6_*,α*_8_*,α*_10_].

**Fig 4.**
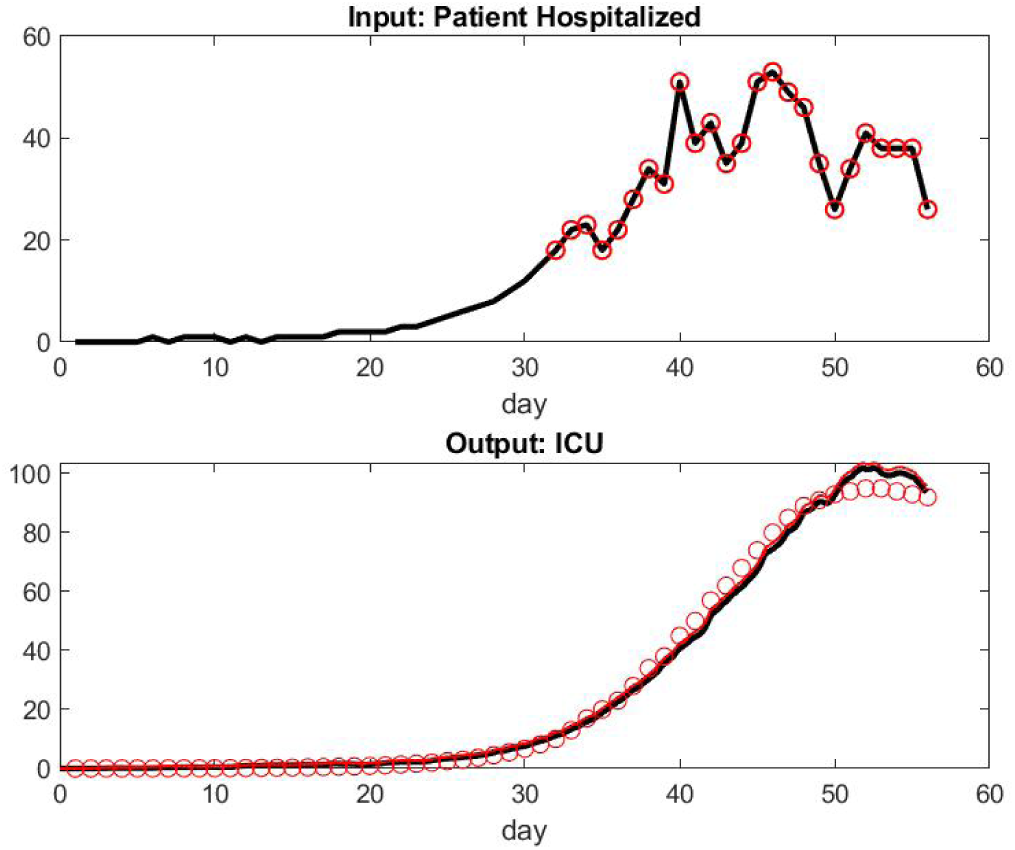
Top: Input of Patient Hospital Admission In French Hospitals. Day 32 corresponds to the first date of our data set i.e 3/18/20. We assume an exponential model for the period anterior to this date. Day 52 corresponds to 4/11/20. Bottom: Model of Patient Under Mechanical Ventilation in ICU versus data set.

**Fig 5.**
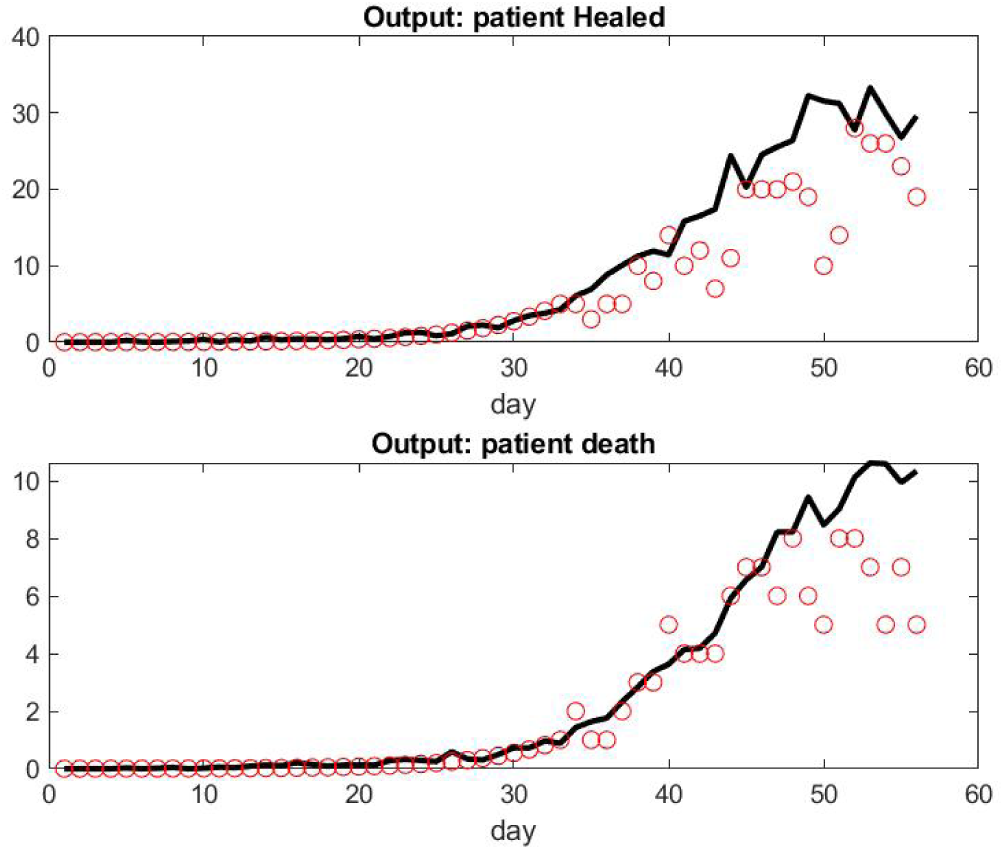
Top: Number of Patient Healed Leaving Hospital per Day. Bottom: Number of Death per Day.

We observed that the number of patient admissions is not a smooth curve. Typically, Sunday’s have less activity with less patients discharged than weekdays. However, the model fitting seems adequate and robust to a small variation of parameters. The logic on the influence of parameters is simple, *α*_2_ being the one who is most important for all output. Each of the six parameters seems to have some significant influence for at least one of the three outputs.

This is not completely surprising because the construction of the model was subject to a number of experimentations and trial errors by way of simulations before getting what we found: a model with low complexity that makes the fitting feasible.

The curve of the number of patients under mechanical ventilation is smooth as expected because this care unit has by far the longest LOS. Consequently, the system has a lot of delays and the relatively small number of patients per day who have been healed or died is still relatively small. Similarly, the number of patients who are mechanically ventilated does not directly reflect the number of admissions at the early stages of the pandemic in the FDS.

For some unknown reasons to us, the model overestimates the number of patient deaths in the last few days. Those numbers might be small however and more sensitive to singular events or simply involve delay in reporting. It might also be the accumulation effect of the small lag differences of the prediction of patients under mechanical ventilation and the reality.

In order to compare the results obtained with designated subset of the FDS that corresponds to the hospital in Paris and the hospitals in Alsace, we used the simulation with the exact same set of parameters found for the data set with the whole country. Alsace has been the busiest cluster at the beginning of the pandemic, followed later on by Paris and Ile de France. The results for Alsace are reported in Figure 6 and Figure 7. We observed a fairly large difference of the model’s prediction on the number of patients under mechanical ventilation. It seems that at the peak of the pandemic in Alsace, the number of patients under mechanical ventilation was less in reality than in the model. One possible factor would be the shortage of available beds in the ICU. On the other hand, the number of deaths did not go higher than significantly expected. A better explanation might be the fact that a fairly large number of patients in critical condition were transferred to hospitals in different parts of the country or neighboring countries: according to local newspaper more than 110 patients from Alsace have been transferred [33]. This seems coherent with our results: the scaling factor for the Alsace data set to get a maximum hospitalization rate of about 50 patients per day is 6; the overshoot on the ICU prediction is about 20 in Figure 6; The total maximum overshoot is therefore about 120; considering that the average LOS in ICU (see Table 1) is roughly 12 days, our model still seems to give an adequate approximation. But unfortunately, we do not have enough information to add this new patient path in the workflow of Figure 1.

**Fig 6.**
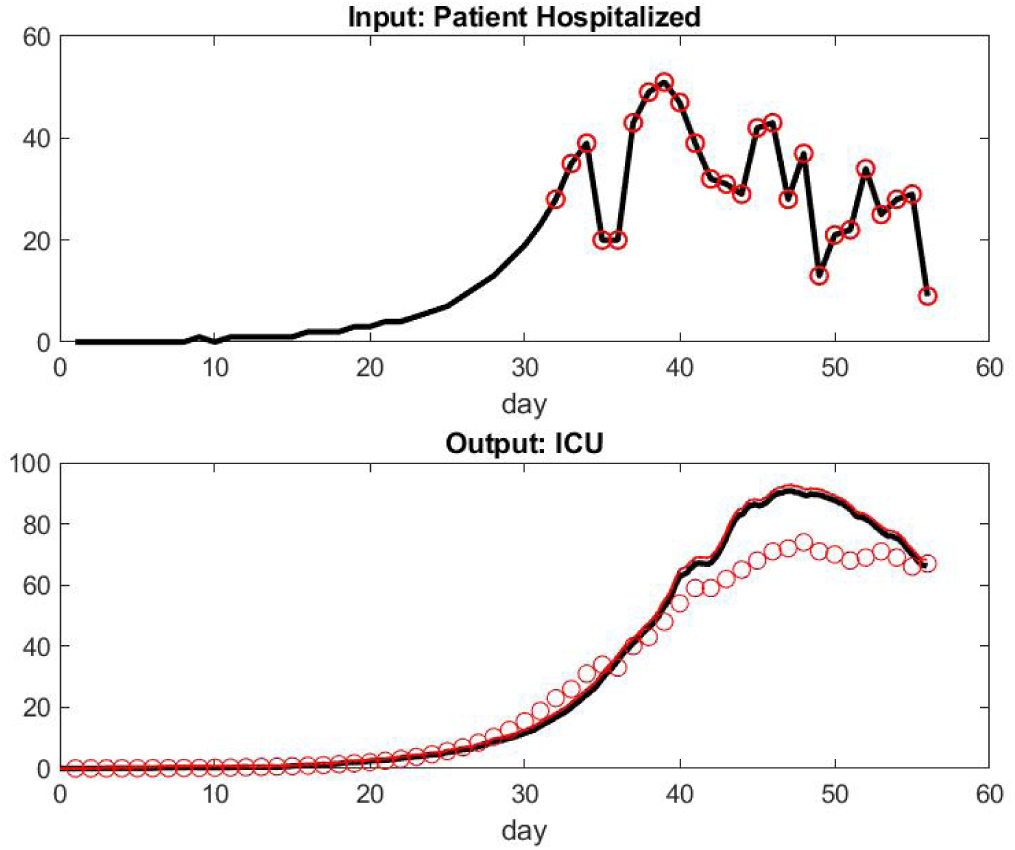
Top: Input of Patient Hospital Admission in Alsace, France. Bottom: Model of Patient Under Mechanical Ventilatin in ICU Versus Data Set.

**Fig 7.**
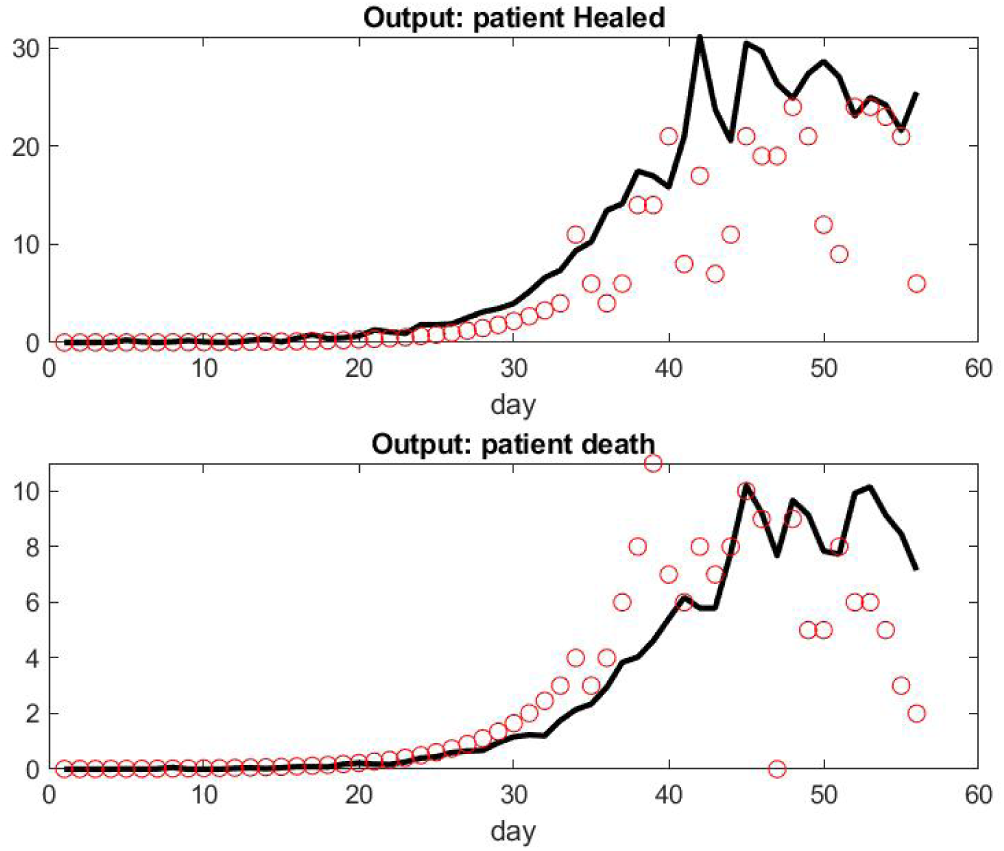
Top: Number of Patient Healed Leaving Hospital in Alsace, France per Day. Bottom: Number of Death per Day.

This phenomena is less present in the results for the data set with Paris but are still there -see Figure 8 and Figure 9. One can indeed refine the parameter fitting to be specific for Alsace and Paris in order to reflect that the clinical decision process in the workflow, i.e parameters of Table 1 to 3, might be sensitive to how much the local system is under stress, but we should then take into account those number of transferred patients that are not negligible.

**Fig 8.**
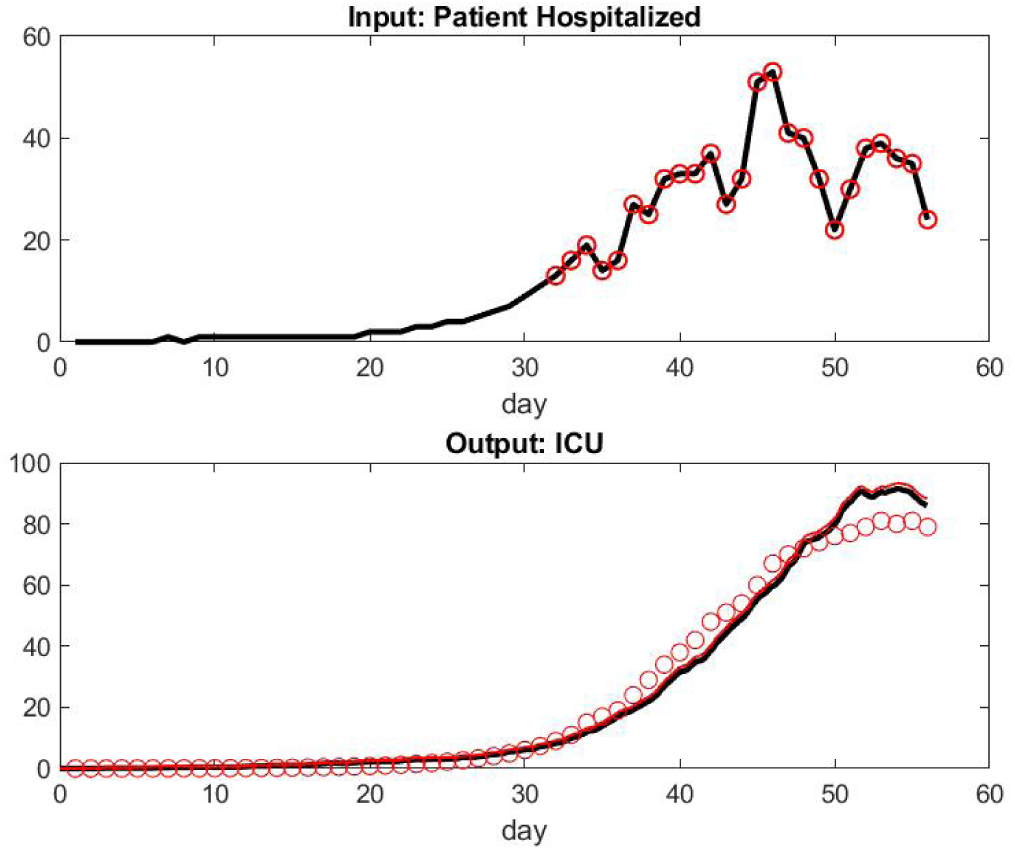
Top: Input of Patient Hospital Admission in Paris, France. Bottom: Model of Patient Under Mechanical Ventilation in ICU Versus Data Set.

**Fig 9.**
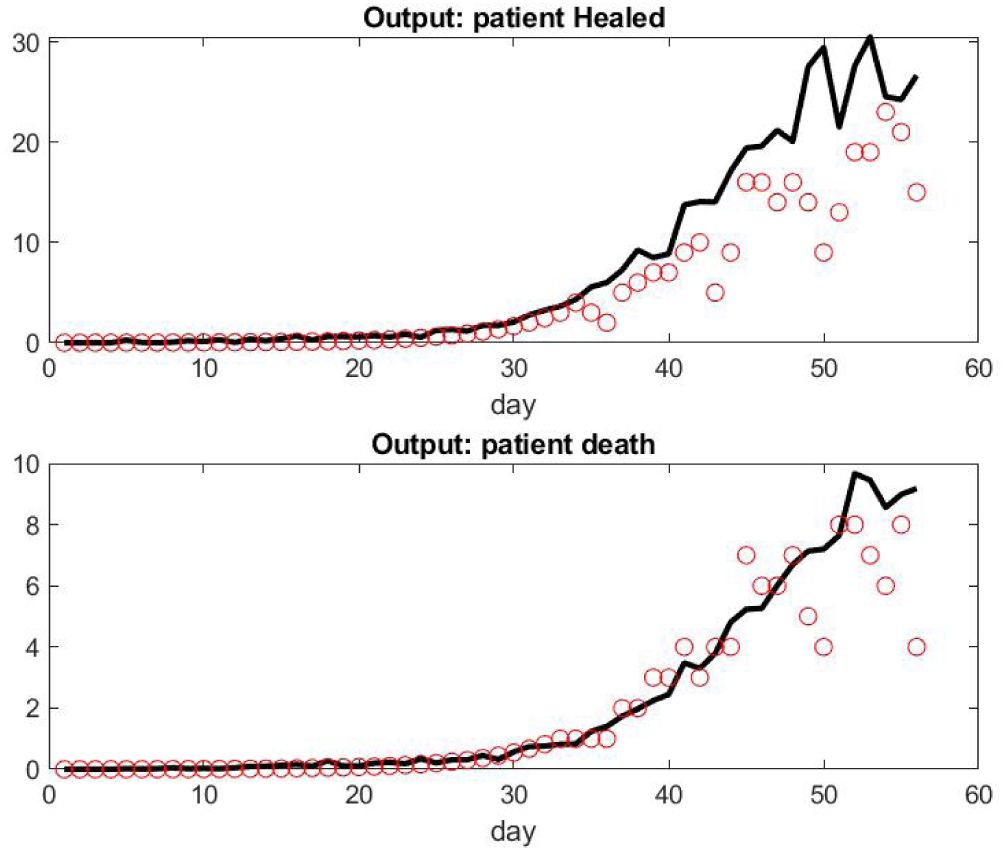
Top: Number of Patient Healed Leaving Hospital in Paris per Day. Bottom: Number of Death per Day.

Next, let us describe the use of our model to assist daily management in the hospital during the pandemic. One key factor is to anticipate the load of each care unit and required resources, either to match the increase in number of patients or to reallocate resources to other patients who have seen their surgery postponed.

We choose a hypothetical scenario that might occur if confinement conditions to contain the pandemic are lifted too early. We assume that the hospital has a nominal low flux of patients from week 1 to 7, and a recurrence with a daily 20% increase of new patients coming in occurs in week 8. Figure 10 shows the dynamic of the load of each care unit, in particular the large delay in the number of patients in the ICU that becomes saturated the latest. The black curves are a simulation of the previous week’s load (week 7), while red curves are the prediction for the following week (week 8). The upper thin red curve shows the deviation up one standard deviation to give a sense of the uncertainty of the estimate. This uncertainty grows as the time of the prediction gets further away. The cyan curve shows an hypothetical capacity of each care unit: the floor gets saturated first and needs new beds after a few days.

**Fig 10.**
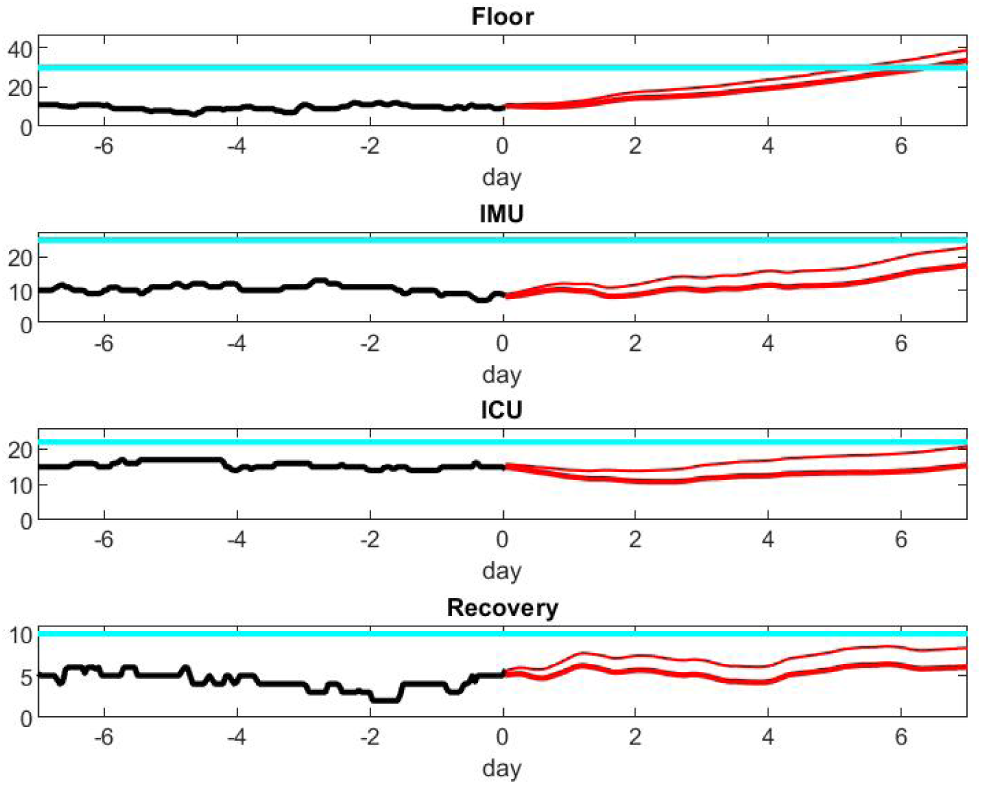
Prediction of Unit Load in Scenario 1.

As an illustration of the capability of the model, Figure 10 and Figure 11 provide an estimate of the growth of resources needed to face the new patient wave. A number of decisions should be made in regards to patient care. Figure 12 compares the patient output with or without shortage of nurses. Those results are speculative since it is difficult to quantify the risk for patients beyond the nice publication results of [9] and [8]. It is our hope that data accumulated during crises such as the present episode of COVID-19 will give the mathematical modeling the base to do this estimate rigorously in future work.

**Fig 11.**
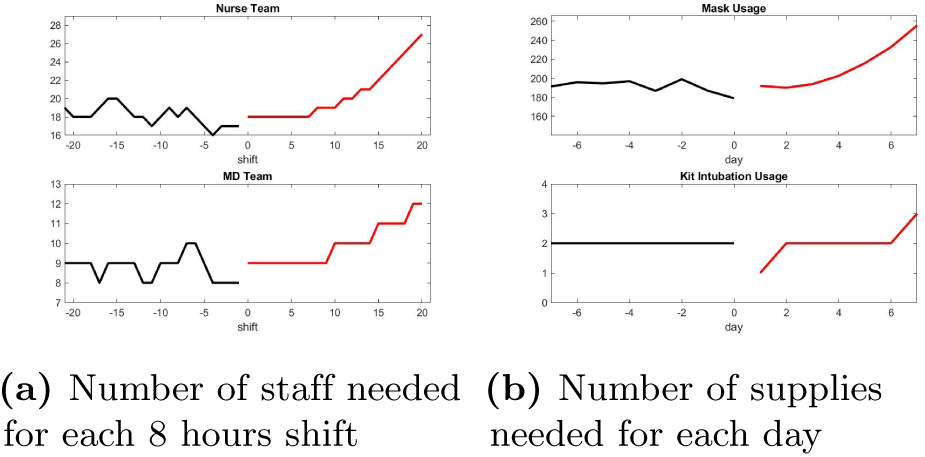
Prediction of Infrastructure Need in Scenario 1.

**Fig 12.**
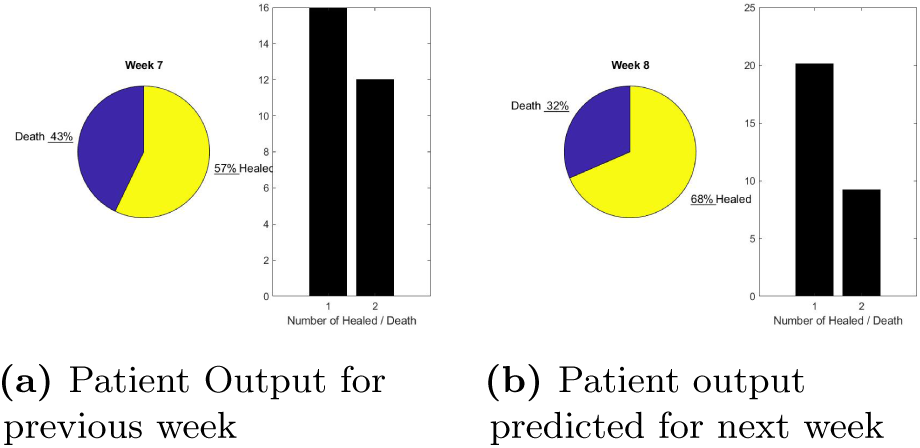
Prediction of Performance in Scenario 1.

Finally taking a step further, we have looked at the prediction of the model to check either the effect of confinement policy on pandemics or new pandemics with an arbitrary rate of transmission of the disease. There are many epidemiological mathematical models available, even for the present crisis, see [25] and [26]. It might be difficult to assess the basic reproduction number *R*_0_ factor, which is under active debate. It is probably even more difficult to assess the exact impact of global confinement or targeted confinement on those parameters that characterized the pandemic model.

We should however be able to use our model to test if the effect on the most critical resource, such as ICU beds and delay in care, are linearly or nonlinearly related to those parameters.

Let us use the most simplistic ordinary differential equation epidemiology model:

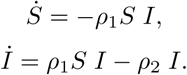

where S is the population of susceptible individuals, and I is the population of infected individuals. *ρ*_1_ is the transmission rate of the disease, and *ρ*_2_ represents the addition of the rate of recovery over infection and the disease induced death.

The function *I*(*t*) is used as the input of our workflow model, and represents the number of patients admitted to the hospital. We test the influence of the transmission rate on the number of ICU beds over a 16-week period. Figure 13 shows that the maximum number of ICU beds required during the epidemic is significantly higher when the transmission rate increases from 0.0015 to 0.0025, while the average number of ICU beds stays about the same. This clearly shows the nonlinear nature of the ICU load management problem and the benefit of a confinement method that lowers the transmission rate during a pandemic.

**Fig 13.**
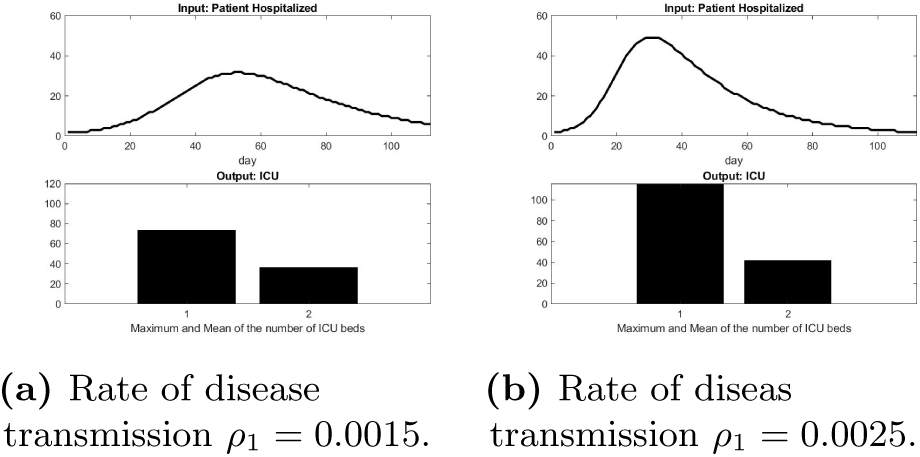
Influence of the disease transmission rate on the number of ICU beds.

## Discussion and Conclusion

In this work, we have developed a simple computational model to mimic the workflow of an average hospital during a pandemic crisis, such as COVID-19 where patient admission goes up to 50 patients per day. This is a significant load for any hospital system because all patients suffer from the same disease and cannot be triaged using the existing departmental structure. The hospital system needs to recruit resources quickly enough to deliver quality patient care while keeping the staff safe from infection.

There are many ways of developing such a mathematical model. We chose a Markov process that can augment a workflow graph provided by the clinicians and used a simple statistical model for the LOS of the patient at each stage corresponding to a graph node. A number of variations in the model construction are available: for example, changing the probability distribution of LOS for specific stages with a more sophisticated model than lognormal or decomposing the graph nodes into subgraphs of the workflow with more details. In particular, the ICU supports different paths of medical care depending on patient conditions. Because of the sparsity of data on hand, we kept the model as simple as possible and we were able to fit the French Data Set with good accuracy. Using this approach, we could:

- recover important parameters that are characteristics of the workflow such as the probability for a patient to transition from one unit to another, and important patient outcomes such as healing rate or death rate.
- on a pragmatic side, we use the model to assist the senior manager in answering his/her questions as listed in our introduction: how many beds do I need on the floor, how is this affecting patient outcomes, do we need to transfer patients to a different facility, etc.?

There are a number of limitations to our approach. The smaller the hospital, the less predictable the outcome will be. With time, the characteristics of the population of patients who show up to the ER may change and the pandemic management by the governing organizations would evolve. One can think, for example, that systematic testing would provide early diagnostics and impact the performance of the health system as shown by the statistics of countries who were early adopters of that strategy. Due to the heterogeneity of the patient population and disease patterns that depend heavily on patient characteristics, our next step in improving this model would be to include patients’ medical history listed in the electronic medical record.

Above all, any model of workflow especially during a pandemic should be aware of the *Human Factor*. Staff can get sick or burnout during a pandemic and there should be a number of strategies to compute that risk and enter this into the constraints imposed on the health care system [4, 11, 12, 21]. Further, human behavior and decision process changes under stress: it can be for economical or psychological reasons. The future of computational models in digital health during a pandemic crisis should extensively include sociological and economical modeling components in the matter.

## Data Availability

All data used in this paper are made available by the French government on their website.

https://www.data.gouv.fr/fr/datasets/donnees-hospitalieres-relatives-a-lepidemie-de-covid-19/#_

https://dashboard.covid19.data.gouv.fr/

## Acknowledgments

*We would like to thank Patrick Doolan for sharing his view with us on management and risk evaluation from his great experience acquired from the energy sector*.

Declarations of interest: none

